# Clinical-grade whole genome sequencing of colorectal cancer and 3’ transcriptome analysis demonstrate targetable alterations in the majority of patients

**DOI:** 10.1101/2020.04.26.20080887

**Authors:** Agata Stodolna, Miao He, Mahesh Vasipalli, Zoya Kingsbury, Jennifer Becq, Joanne D Stockton, Mark P Dilworth, Jonathan James, Toju Sillo, Daniel Blakeway, Stephen T Ward, Tariq Ismail, Mark T. Ross, Andrew D. Beggs

## Abstract

**Introduction:** Clinical grade whole genome sequencing (cWGS) has the potential to become standard of care within the clinic because of its breadth of coverage and lack of bias towards certain regions of the genome. Colorectal cancer presents a difficult treatment paradigm, with over 40% of patients presenting at diagnosis with metastatic disease. We hypothesised that cWGS coupled with 3’ transcriptome analysis would give new insights into colorectal cancer.

**Methods:** Patients underwent PCR-free whole genome sequencing and alignment and variant calling using a standardised pipeline to output SNVs, indels, SVs and CNAs. Additional insights into mutational signatures and tumour biology were gained by the use of 3’ RNAseq.

**Results:** Fifty-four patients were studied in total. Driver analysis identified the Wnt pathway gene *APC* as the only consistently mutated driver in colorectal cancer. Alterations in the PI3K/mTOR pathways were seen as previously observed in CRC. Multiple private CNAs, SVs and gene fusions were unique to individual tumours. Approximately 20% of patients had a tumour mutational burden of >10 mutations/Mb of DNA, suggesting suitability for immunotherapy.

**Conclusions:** Clinical whole genome sequencing offers a potential avenue for identification of private genomic variation that may confer sensitivity to targeted agents and offer patients new options for targeted therapies.

## INTRODUCTION

Colorectal cancer (CRC) is one of the most common malignancies, with over 30,000 cases reported in the UK in 2015–2016 and a 5 year survival rate of approximately 60% (1). CRC is typically initiated by a mutation in the Wnt signalling pathway gene *APC* (2) (adenomatous polyposis coli) or associated genes (*CTTNB1, RNF143, RSPO2/3*) that leads to the formation of a polyp (3) that then progresses via mutations in a number of oncogenes and tumour suppressors into an invasive cancer. In parallel with the expansion of our knowledge of the biology of colorectal cancer, the field of targeted oncology is rapidly advancing, with targeted agents available (4) for a high percentage of driver and modifier mutations across a wide range of cancers.

The Cancer Genome Atlas (TCGA) project set out to characterise mutations in colorectal cancer by exome sequencing of a cohort of 600 patients using the Agilent SureSelect panel via tumour-normal subtraction (5). It confirmed recurrent mutations in *APC, TP53, SMAD4, PIK3CA* and *KRAS* as well as identifying recurrent mutations in *ARID1A, SOX9* and *AMER1* (*FAM123B*). Giannakis *et al*. (6) carried out exome sequencing on a clinically annotated cohort of 619 patients, finding further recurrent mutations in *BCL9L, RBM10, CTCF* and *KLF5*, and also showing that neoantigen load as determined by exome sequencing was associated both with tumour associated lymphocyte infiltration and overall survival. However, a key weakness of the TCGA and other studies has been the use of exome sequencing to demonstrate key oncogenic drivers. Exome sequencing, whether by the amplicon or hybridisation approach, may miss key oncogenic drivers due to allelic drop out or the biases inherent to targeting approaches (7).

Whole genome sequencing has a number of potential advantages. Firstly, it can increase overall variant calling accuracy as exome sequencing techniques can suffer from probe drop out and poor coverage, especially at splice junctions and in "difficult” to sequence regions where probe drop out is common (8). Secondly, it can natively call fusions (9) and other structural variants (10) (by detection of split reads); and finally it can identify copy number variants (11) to a higher accuracy than alternative techniques. Given the recent attention to tumour mutational burden (TMB) in selecting patients for anti-PD1 therapies such as pembroluzimab, whole genome sequencing can accurately call mutation burden (12).

However, until very recently, studies of colorectal cancer using whole genome sequencing have been limited in number or scope. Shanmugan *et al*. (13) carried out whole genome sequencing in order to identify therapeutic targets in four patients with metastatic disease, finding several known mutations of interest as potentially targetable. Ishaque *et al*. (14) carried out paired metastasis-primary tumour whole genome sequencing in colorectal cancer, finding novel non-coding oncogenic drivers and an elevated level of “BRCAness”. The Pan Cancer analysis of whole genomes (PCAWG) consortium (15) presented 52 colorectal (37 colon, 15 rectal) whole genome sequenced tumours as part of the larger consortium effort, although at the time of writing, no specific examination of the landscape of these had been carried out, presumably because of the previous TCGA colorectal cancer paper which examined the exomes of 276 colorectal cancers (5).

The United Kingdom 100,000 Genomes project has set out to sequence tens of thousands of cancer genomes (16), across multiple tumour types, using a clinical grade sequencing pipeline and variant calling algorithm. Our study has carried out whole genome sequencing of 54 paired colorectal tumour-normal samples, utilising a the Genomics England clinical-grade sequencing, alignment, variant calling and annotation pipeline in order to understand the utility of WGS in colorectal cancer.

## METHODS

### Patients

Sequential patients undergoing elective colorectal surgery at the Queen Elizabeth Hospital Birmingham were recruited for the study. Patients were selected who had sporadic colorectal cancer and did not have an Amsterdam positive history of colorectal cancer or an age of onset less than 45 years. Consent for the study was taken and the study was fully ethically approved by the University of Birmingham Human Biomaterials Resource Centre (HBRC, ethical approval ref 15/NW/0079).

### Samples

Immediately after resection, resected specimens were conveyed to a histopathologist who facilitated direct biopsy of tumour material and associated normal bowel (defined as the distal resection margin) by frozen section. Samples were immediately snap frozen on liquid nitrogen and stored at −80°C until needed. Tumour content was verified by frozen section, with at least 60% tumour being needed for inclusion in the study. DNA was extracted using a Qiagen DNEasy kit and RNA with a Qiagen RNEasy kit. Nucleic acid quantity and quality were assessed using a Qubit2 fluorimeter and TapeStation assay.

### Library preparation

Sequencing libraries of 500 ng DNA extracted from fresh-frozen tumour or normal tissue were prepared using the TruSeq® DNA PCR-free method (Illumina). Sequencing (100 base paired reads) was performed on the HiSeq2500 platform to a mean depth of >30x for the normal genome and >60x for the tumour genome, after the removal of duplicate read-pairs.

RNA: Libraries were prepared using 50 ng of RNA using a Lexogen QuantSeq 3’ RNA kit from tumour and matched normal samples. Polyadenylated mRNA was pulled down then cDNA synthesis and 3’ library preparation carried out. Samples were indexed and pooled across an Illumina NextSeq v2 flow cell and sequenced using a 75 base single-ended sequencing strategy

### Bioinformatics

WGS: Raw reads were converted to FASTQ using bcl2fastq, quality trimmed then mapped to the GRCh37 (hg19) Human Reference Genome using the Isaac3 (17) aligner (Illumina). Single nucleotide and indel variants were mapped using the Strelka2 (18) variant caller (for the germline calls using germline-only mode and for somatic calls using joint tumour/normal mode), somatic structural variants using the Manta (19) structural variant caller and copy number aberrations using the Canvas (20) copy number caller. Annotation of the variants was performed using Illumina’s annotation engine Nirvana (https://github.com/Illumina/Nirvana/wiki) using Ensembl 73 as database reference. Novel driver analysis was generated using MutSigCV2(21), Intogen(22) and dNdScv(23) with and without hyper-mutated samples. Non-coding driver analysis was performed with FunSeq2 (24). Mutational Signatures were generated using the MutationalPatterns R/Bioconductor package (25). All variants were stored in VCF files. Telomere length from whole genome sequencing data was derived using TelomereCat (26)

Copy number calls were pooled across individuals with bedtools and overlapped with bedIntersect to identify regions that were recurrently gained/lost. Structural variants were pooled using bedtools and overlapped with intersectBed to identify common regions of structural variation.

### In samples requiring mutational confirmation, Sanger sequencing was performed (primer sequences available on request)

RNA-seq: FASTQ files were quality trimmed, adapters were removed and reads were aligned to the hg19 reference genome using the STAR aligner (27) (version 2.6.1). Genes were annotated using the Ensembl v74 database and gene-centric read counts generated using Partek Flow GSA algorithm (28). Hierarchical clustering and PCA plots were also generated. CMS and CRIS signatures were called using the CMSCaller R package (29). For calculation of the CIRC score, the methodology of Lal *et al*. was used (30). For immune infiltration scores via CIBERSORT, the methodology described by Chen *et al*. was used (31). For the signature derivation, the BioSigner module of Bioconductor was used (32).

### Data availability

All data are available in the European Nucleotide Archive (accession number XX)

## RESULTS

### Sequencing metrics

In total, 54 tumour-normal pairs (30/54 male, 24/54 female) underwent whole genome sequencing, with a median read depth of 68x for tumour samples and 38x for normal samples. Median purity based on WGS data was 68% (range 29-100%). Median somatic SNVs were 19,700 (range 2,459-1,601,093), somatic indels 4,231 (range 360-464,252) and SVs 105 (range 6-681). Median chromosome count was 46.5 chromosomes/genome (range 41-67). Median tumour mutational burden was 8.04 mutations/Mb (range 0.92-577.91 mutations/Mb).

### Clinical data

In the patients studied, all had primary colorectal cancer. Two patients with rectal cancer underwent neoadjuvant chemoradiotherapy and one underwent neoadjuvant short course radiotherapy before excision of the primary tumour. The pathological stage of the resected tumours varied from between T2N0 to T4N2. Five patients presented with metastatic disease and 18 patients had “high-risk” disease consisting of any of poor differentiation (4 patients), extra-mural vascular invasion (18 patients) or threatened circumferential resection margin (2 patients). The operation types were abdomino-perineal excision of rectum (1 patient), anterior resection of rectum (25/54), left hemicolectomy (5/54), panproctocolectomy (1 patient), right hemicolectomy (16/54), sigmoid colectomy (4/54) and subtotal colectomy (2/54). Median numbers of lymph nodes identified by histopathological examination was 24 (IQR 18-28).

Fifteen patients underwent adjuvant therapy consisting of capecitabine (1 patient), or capecitabine and oxaliplatin (14 patients). Seventeen patients had disease recurrence, with a median time to recurrence of 639 days (IQR 276-2501 days). Fourteen (25.9%) patients died whilst within the study, with a median time to death of 598 days (IQR 398–1231 days).

### Germline mutations

The germline genome of all patients was studied for mutations in genes associated with familial colorectal cancer syndromes (*APC, MYH, MLH1, MSH2, MSH6, PMS2, POLE, POLD1, SMAD4, and BMPRA1*). We found no SNVs or indel germline mutations in this cohort of patients.

### Hypermutator phenotype

In total, 17/54 patients (table 1) had greater than 10 somatic mutations per megabase, suggesting that they may be suitable for anti-PD1 immunotherapy. Of these patients, five had somatic mutations that have previously been demonstrated as responsible for hypermutated tumours (table 1). One tumour had a *POLD1* (p.Leu227Pro) mutation, with a TMB of 206.26 mutations/Mb and a second had a *POLE1* (p.Pro286Arg) mutation, with a TMB of 577.91 mutations/Mb. The other three patients had variants in the mismatch repair genes *PMS1* and *MSH3* (TMB 41.1, 71.1 and 45.2 muts/mb). A further patient had a TMB of 143.31 mutations/Mb with no obvious germline or somatic mutation causing this phenotype.

**Table 1.** –Hypermutated samples with TMB > 10 and potential somatic variants known to be associated with hypermutation.

| Sample | TMB (Muts/mb) | Potential somatic variants |
| --- | --- | --- |
| A03 | 206.25 | POLD1 (p.Leu227Pro) |
| A09 | 85.47 | PMS1 (p.Ser118Ter) |
| A10 | 95.41 | MSH3 (p.Val393Ala) |
| A12 | 143.31 | None detected |
| B05 | 221.68 | MSH3<br>(p.Lys383ArgfsTer32)<br>MLH3 (p.Lys383ArgfsTer32)<br>POLE (p.Arg759Cys) |
| B08 | 577.91 | POLE (p.Pro286Arg) |

### Most frequently mutated genes and identification of new drivers

A generic analysis of the ten most frequently mutated genes (both SNV and indel, not normalised by transcript length) demonstrated that these were (from most to least recurrent): *TTN, APC, MUC4, FAT2, TP53, FRG1, KRAS, LRP2, CSMD3* and *MT-ND4* (Figure 1).

**Figure 1:**
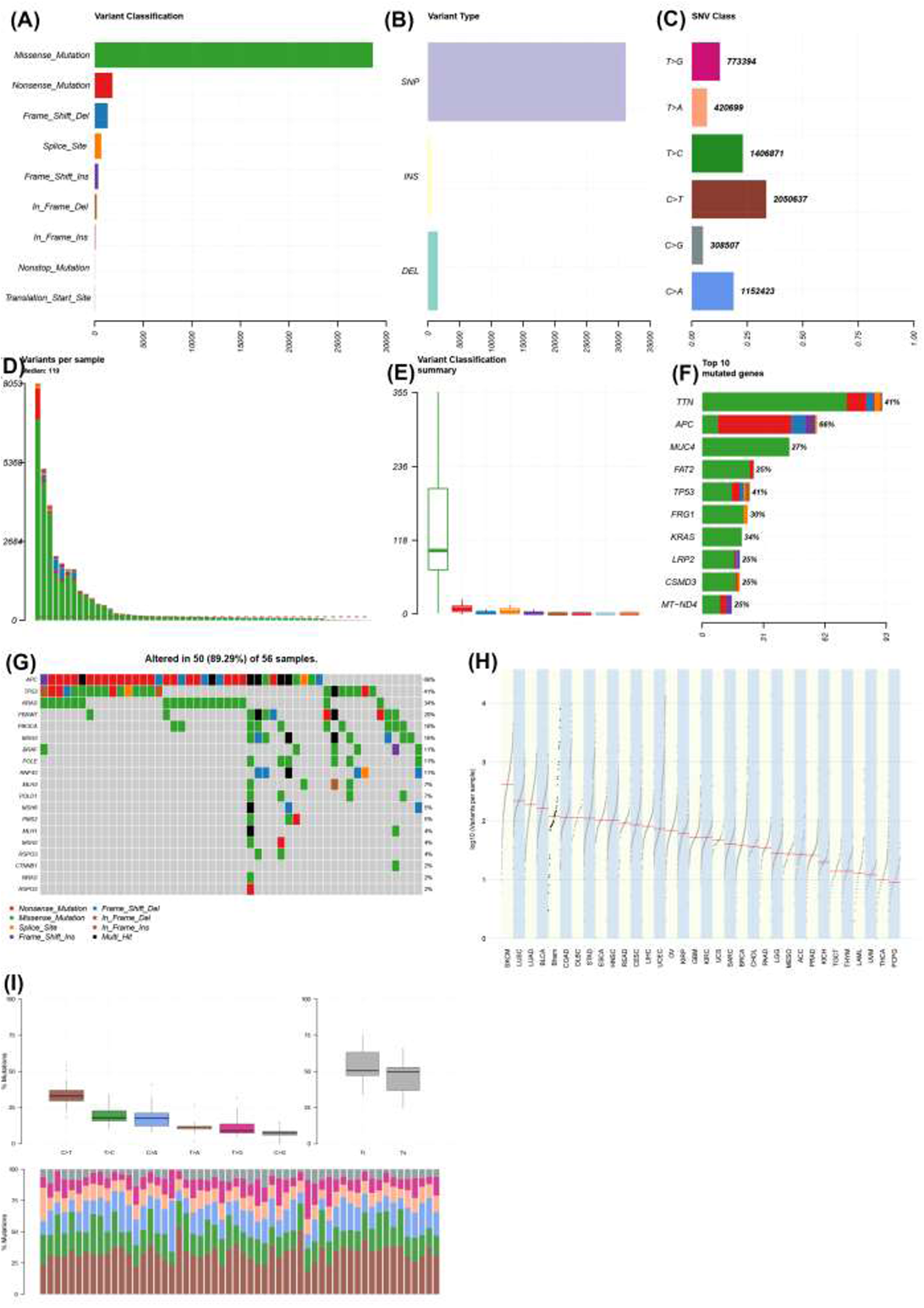
Integrated plot of characteristics of whole genome sequencing dataset of colorectal cancer. A- Variant classification by type (Y-axis), Frequency of variant (X-axis) B- Variant type (Y-axis), SNP = single nucleotide polymorphism, INS = insertion, DEL = deletion; frequency (X-axis) C- Single nucleotide variant (SNV) class plot - Y-axis demonstrates nucleotide changes. X-axis demonstrates proportions of variants in cohort; numbers on end of bars demonstrate total numbers of each variant D- Bar chart showing variants per sample – variants (y-axis); samples on x-axis E- Variant classification summary showing range of variants per sample (y-axis), x-axis shows missense (green), nonsense (red), frame shift deletion (blue), splice site (yellow), frame shift insertion (purple), in frame deletion (brown), in frame insertion (dark red), non-stop mutation (light blue), transcription start site mutation (orange) F- Top ten mutated genes by frequency – genes on y-axis, numbers of mutations on X-axis; colours the same as panel E G- Oncoprint of colorectal driver genes (left Y-axis) by sample (X-axis) with variant type shown in key. Percentages across whole cohort seen in percentages down right Y-axis H- TCGA style log(10) variants per sample plot (y-axis) with TCGA cohorts (X-axis), Bham = Birmingham cohort (fifth from left) I- Mutational type plot: Top left panel - % mutation changes in cohort Top right panel - % transition vs. transversion mutations across cohort Bottom panel – Bar chart showing proportion of mutations with percentage on y-axis and type of mutations shown by different coloured bars

Mutational frequency of cancer genes was compared to known cancer drivers in (Figure 1). The most frequently mutated gene was *APC* (38/54 samples), followed by *TP53* (23/54 samples), *KRAS* (19/54 samples) and *FBXW7* (12/54 samples). Less frequent mutations were seen in genes that are typically considered “druggable” but not seen previously in colorectal cancer including *KIT, ERBB2* and *ALK*.

For all driver analyses, samples were analysed in hypermutated and nonhypermutated groups. For the hypermutated analysis, MutSigCV analysis (in order to identify genes significantly mutated compared to background) of driver mutations demonstrated 1,235 potentially significant mutations (p<0.05, q<0.05) in the dataset. Only *APC* was highlighted as significant from the typical colorectal driver mutations (Supplementary table 1). For the non-hypermutated analysis, MutSigCV analysis demonstrated 97 potentially significant mutations, with *APC, TP53, KRAS, SOX9* and *FBXW7* being highlighted as significant driver genes (Supplementary table1).

Intogen analysis (in order to identify genes under positive selection) of the hypermutated set (Supplementary table 2) revealed 80 genes as potential drivers via either OncoDriveFM or OncoDriveClust. The top five drivers as determined by order of significance were *APC* (P_oncodriveFM_=0, Q_oncodriveFM_=0), *TP53* (P_oncodriveFM_=0, Q_oncodriveFM_=0), *KMT2C* (P_oncodriveFM_=6.56×10–4, Q_oncodriveFM_=0.042), *KRAS* (P_oncodriveFM_=3.11E-15,Q_oncodriveFM_=03.35E-12) and *HLA-A* (P_oncodriveFM_=5.43E-1 0, Q_oncodriveFM_=4.88E-07). In the non-hypermutated set, 333 genes were flagged as potential drivers with the top 5 being: *APC* (P_oncodriveFM_=0, Q_oncodriveFM_=0), *TP53* (P_oncodriveFM_=0, Q_oncodriveFM_=0), *KRAS* (PoncodriveFM^=^4.44E-16,Q_oncodriveFM_^=^03.93E-13), *SOX9* (P_oncodriveFM_-7.16E-14,Q_oncodriveFM_=4.75E-11) and *FBXW7* (P_oncodriveFM_-1.38E-13, Q_oncodriveFM_=7.33E-11).

dNdScv analysis (in order to identify genes under positive selection) of the hypermutated set (Supplementary table 3) demonstrated 10 genes with p<0.05 and Q<0.1, the top ranked one of which was *FRG1*, followed by *KRAS, TP53, APC, DYNC1I2, FBXW7, AC093323.1, PIK3CA, IGSF3* and *PTEN*. For the nonhypermutated set 5 genes had p<0.05 and Q<0.1, the top ranked one being *FRG1* followed by *KRAS, APC, TP53* and *SOX9* (Supplementary table 3).

An analysis of non-coding drivers using FunSeq2 (33) revealed multiple regions with statistically significant increased mutation rates as compared to background (supplementary table 4). In the hypermutated set, the top ranked region (Chr2:133021792-133036207) was identified as having recurrent mutations and is predicted *in silico* to bind the BRCA1, CHD2, IRF3, MAFK, MXI1, NFKB1, RFX5 and SMC3 transcription factors. The long non-coding RNA ENSG00000232274.1 (chr1:143,189,434-143,211,070) was also recurrently mutated. In the nonhypermutated sample set, the AP-1 transcription factor complex member JUND was recurrently mutated in 46/47 samples in non-coding regions. FunSeq2 analysis also ranked *APC* as the top ranked coding driver mutation in 27/54 samples.

An overlapping analysis of potential drivers using Venny from all four algorithms only demonstrated *APC* as being a potential driver in the dataset across all four sets of calls (Figure 2). When the MutSigCV calls were removed, 12 genes were enriched (*KRAS, TP53, FBXW7, PIK3CA, NPEPPS, CTNND1, FLII, MGA, SETPB1, BCL9, MSH3* and *ANXA6*). When the Intogen calls only were removed, 4 genes were enriched (*ZNF517, CROCC, TPO* and *FSHR*). When dNdScV was removed, *RIPK4* only was enriched and when FunSeq2 calls only were removed there were no significant genes.

**Figure 2:**
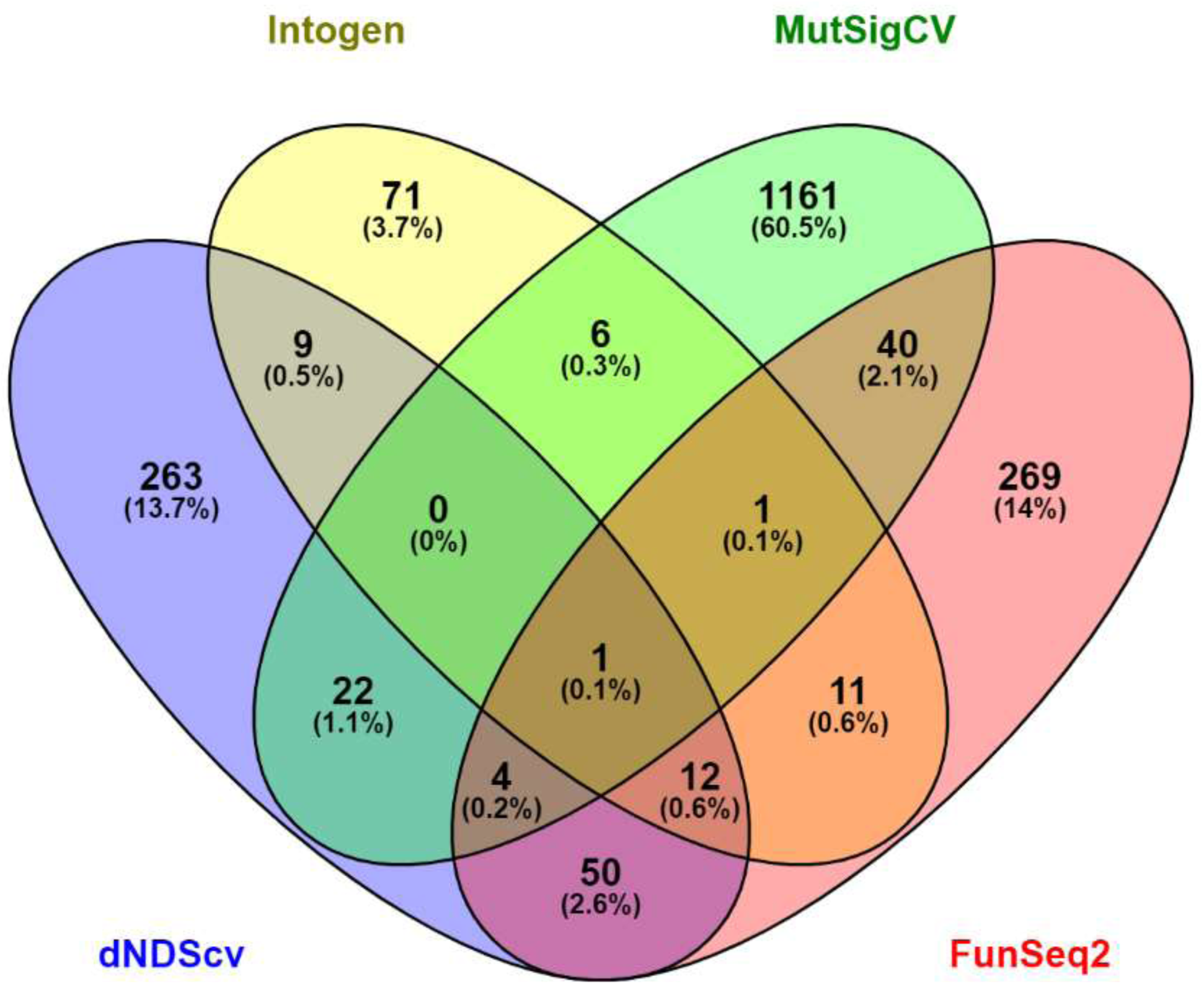
Overlapping genes from each significant variant caller (Intogen, MutSigCV, dnDScv and FunSeq2)

A pathway analysis of these pooled drivers across the four algorithms using G Profiler (34) revealed enrichment in a number of transcription factor associated enriched pathways, KEGG pathways, and GO terms (supplementary table 5)

### Copy number aberrations

A pooled analysis of copy number variation across the cohort was performed (Figure 3). A consistent low-level pattern of both copy number gain and loss was observed. When filtered by exonic regions across all samples, 6/354 losses and 2/30 gains were observed to be exonic. Gains were seen in all samples in the *FOXI2* (chr10:129534543-129537433) and *REX1BD* genes (chr19:18654566-18746304). *FOXI2* is a forkhead binding gene associated with transcriptional activation which has been seen to be consistently hypomethylated in colorectal cancer (35) and *REX1BD* (Required For Excision 1 Binding Domain) is a putative DNA repair gene (36).

**Figure 3:**
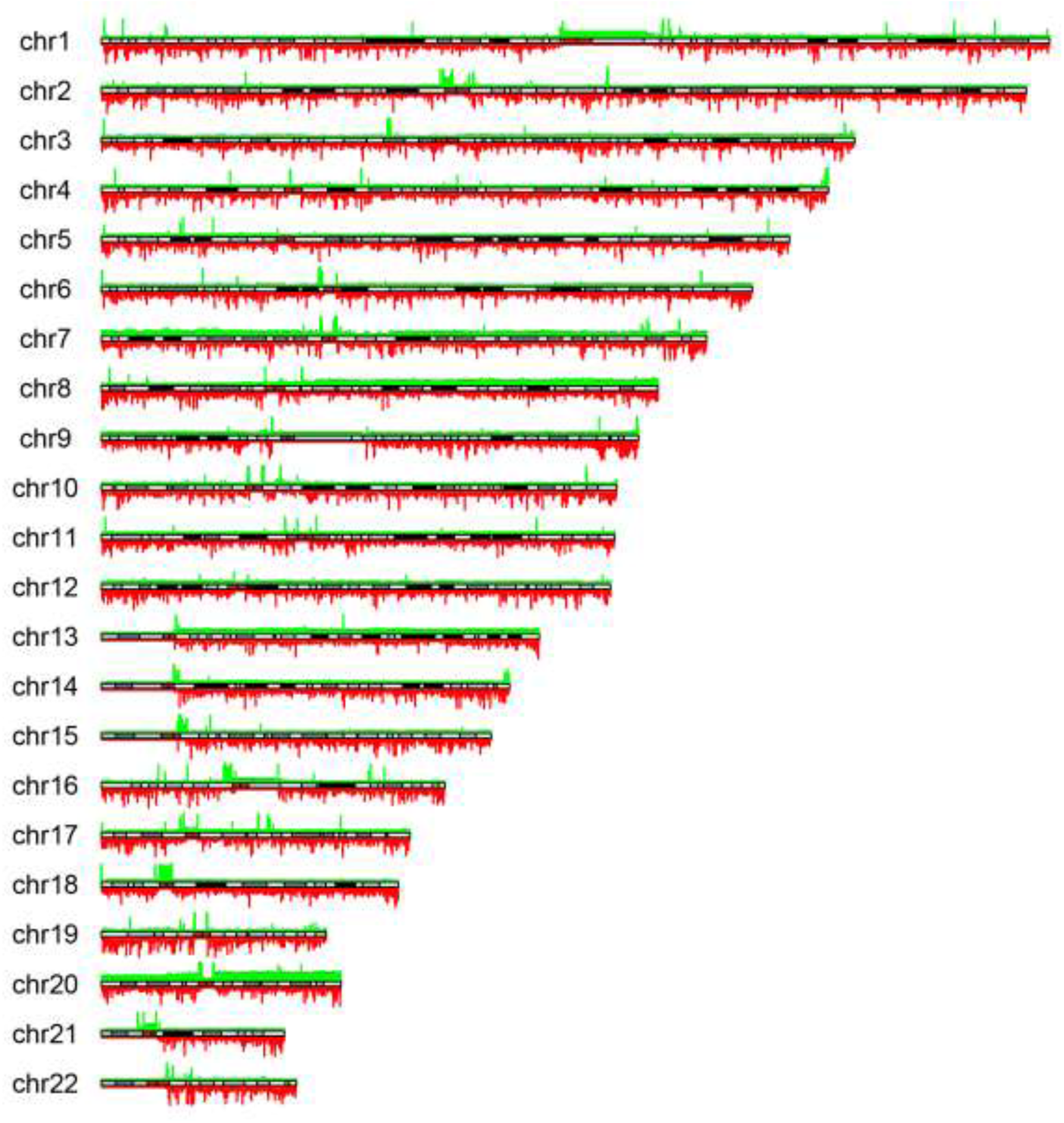
Genome wide copy plot all samples across cohort (green – gain, red – loss); Height of bar is proportional to number of samples with copy number variation.

Losses were seen for all samples in *MYO1C* (chr17:1385365-1386295), *CBARP* (chr19:1230748-1231737), *PIMREG* (chr17:6358505-6359232), *NFATC1* (chr18:77159859-77161091), *UCN3* (chr10:5415602-5416345) and *AMH* (chr19:2247518-2248270). MYO1C controls nuclear membrane tension (37), has been previously reported as recurrently deleted in gastric cancer (38) and is thought to have a role in PIK3 signalling (39). *NFATC1* is a gene of the nuclear factor of activated T cells (NFAT) class, which have been shown to play a key role in the progression of solid tumours (40).

### Structural Variants

Structural variants were filtered on the basis that the most functionally relevant ones were likely to be those involving known cancer driver genes. In total, 29 potential oncogenic gene fusions, detected by WGS were seen in 16 samples (Table 2). Of the 29 potential gene fusions, no recurrent gene fusions were seen. However, fusions involving *IDH1-PTH2R, CDK6-CDK14, KAT6B-RBMS3, ERBB2-HAP1, CCDC6-TMEM212AS1* and *BRAF-DLG1* were seen.

**Table 2:** List of potentially oncogenic structural variants in cohort.

| Gene | Consequence | Chromosome | Position | Ref | Alt |
| --- | --- | --- | --- | --- | --- |
| GNAM11 | GNA11 i > HNRNPM i > | chr19 | 3109401 | intron | DEL |
| NRG1 | NRG1 i > L3HYPDH i > | chr8 | 32154965 | intron | BND |
| SMAD4 | MRO i < SMAD4 i < | chr18 | 51065790 | intron | INV |
| PTPRK | MAN1A1 i > PTPRK i > | chr6 | 128452884 | intron | DUP |
| IDH1 | VRK2 i < IDH1 i < | chr2 | 208257287 | intron | INV |
| IDH1 | IDH1 e < PTH2R i < | chr2 | 208242100 | exon | INV |
| CDK6 | CDK14 i > CDK6 e > | chr7 | 92612231 | exon | INV |
| CDK6 | CDK14 i > CDK6 e > | chr7 | 92612048 | exon | INV |
| SRGAP3 | LMCD1-AS1 i < SRGAP3 i < | chr3 | 9168820 | intron | DEL |
| NGR1 | LDAH i > NGR1 i > | chr8 | 32654703 | intron | BND |
| NRG1 | LDAH i < NGR1 i < | chr8 | 32654498 | intron | BND |
| KAT6B | RBMS3 i > KAT6B e > | chr10 | 75031261 | exon | BND |
| FAM46C | MAN1A2 i > FAM46C i > | chr1 | 117619648 | intron | DEL |
| SMAD4 | CTIF i < SMAD4 i < | chr18 | 51062229 | intron | DUP |
| RARA | RARA i < TTC25 i < | chr17 | 40354212 | intron | DUP |
| NRG1 | NRG1 i < UNC5D I < | chr8 | 32192673 | intron | DUP |
| CDK12 | FBX047 i < CDK12 i < | chr17 | 39521585 | intron | INV |
| CDK12 | PLXDC1 i > CDK12 i > | chr17 | 39465725 | intron | INV |
| ERBB2 | ERBB2 e > HAP1 e > | chr17 | 39727989 | exon | INV |
| ZNF521 | MCHR2-AS1 i < ZNF521 i < | chr18 | 25327550 | intron | BND |
| PPP6C | SCAI i < PPP6C i < | chr9 | 125174977 | intron | DEL |
| EML4 | EML4 i > MTA3 i > | chr2 | 42284332 | intron | DEL |
| BRD4 | BRD4 i < AKAP8 e < | chr19 | 15332325 | intron | DEL |
| KMT2C | KMT2C i < TPTEP1 i < | chr7 | 1522434118 | intron | BND |
| CCDC6 | TMEM212-AS1 i < CCDC6 e < | chr10 | 59788825 | exon | BND |
| CCDC6 | TMEM212-AS1 i > CCDC6 e > | chr10 | 59906506 | exon | BND |
| BRAF | DLG1 i > BRAF e > | chr7 | 140794385 | exon | BND |
| GPHN | GPHN i > FAM71D i > | chr14 | 66721301 | intron | DEL |
| ELL | RFX2 i < ELL i < | chr19 | 18478021 | intron | DEL |

### Mutational signatures

The top three most frequent mutational signatures (V3 SBS signatures (41)) (Figure 4) as determined at the cohort level using the somatic SNVs of all samples were Signature 1 (53/54 samples), Signature 5 (53/54 samples) and Signature 40 (27/54 samples). Signature 1 is the “Ageing” signature and is associated with the consequences of normal tissue ageing, mainly spontaneous cytosine deamination. Signature 5 is associated with tobacco smoking and Signature 40 is also associated with ageing. Other signatures seen were Signature 44 (defective DNA mismatch repair), 17a (pre-treatment with fluorouracil), 17b (pre-treatment with fluorouracil), 13 (APOBEC), 20 (concurrent POLD1 and MMR deficiency), 4 (direct damage by tobacco smoke), 7c (UV radiation), 9 (IGHV hypermutation), 18 (Reactive oxygen species) and 41 (unknown). Signatures 57, 46, 47 were also seen which are known to be due to sequencing artefact.

**Figure 4.**
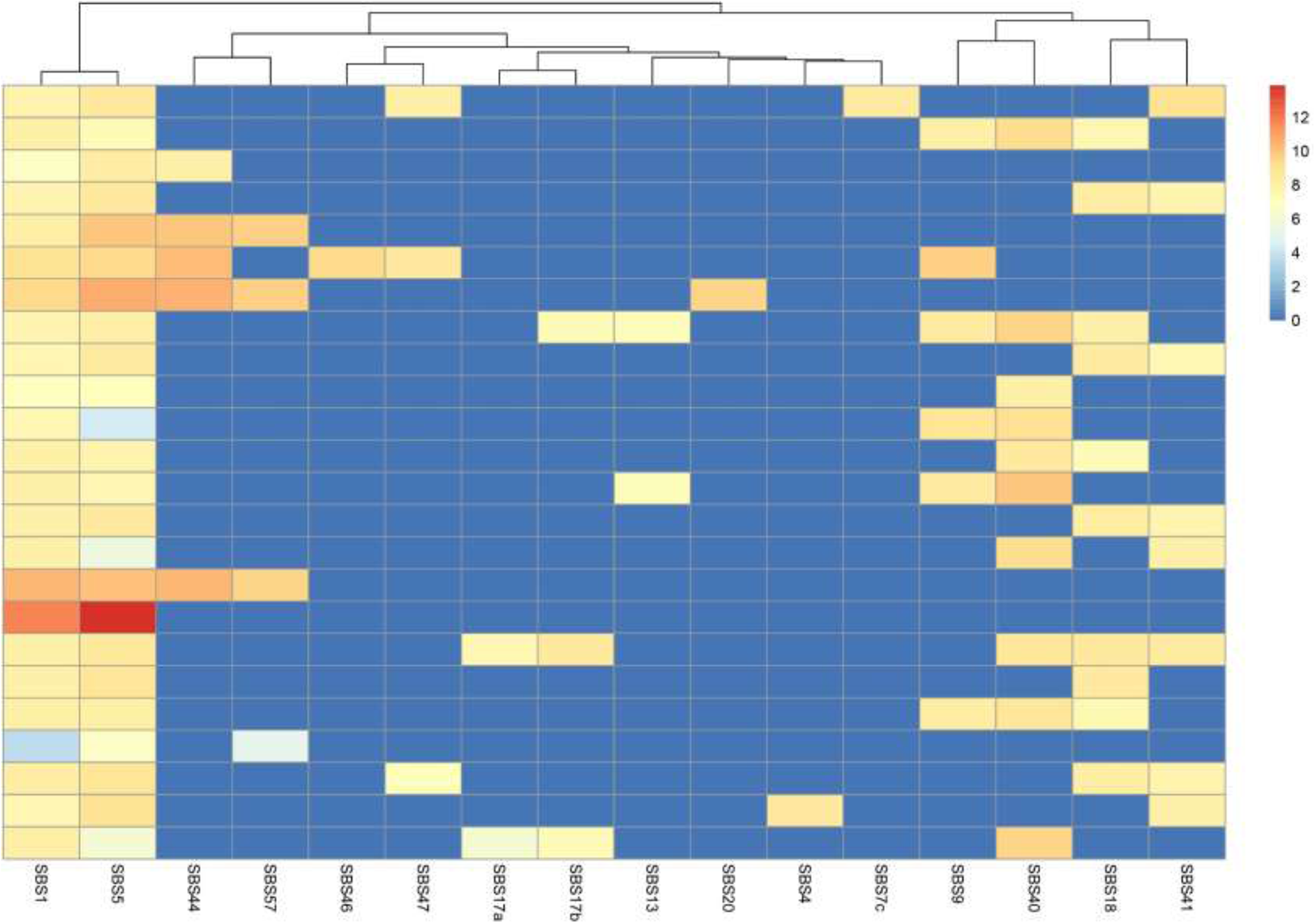
– Most frequent single base substitution mutational signatures shown in hierarchical cluster plot (samples with identical signature combinations were collapsed)

### Kataegis

The phenomenon of kataegis (localised somatic hypermutation) has been previously demonstrated in breast cancer (42). In our study, we found that it occurred in all 54 samples significantly to one extent or another (Supplementary table 1). Kataegis occurred particularly frequently at a per sample level between chr20:31050000-31080000 (Supplementary figure 1) which corresponds to the region of *NOL4L/C20orf112* (chr20:31,030,862-31,071,385) a known fusion partner of *RUNX1* and *PAX5* in leukaemia (43).

### Telomere length

Because of the well observed phenomenon of shorter telomere length in cancer, we studied the lengths of telomeres as measured by whole genome sequencing, which have previously been shown to correlate well to older methods such as Southern blotting (26). Median telomere length in cancer was 5,028 bp and in normal germline blood was 6,294 bp (Mann-Whitney p<0.0001).

## RNA-seq

### Differential expression profiles

In order to understand if there were any *de novo* transcriptional subgroups within the dataset, a cut-off of the top 250 genes by variance was extracted from the dataset. When comparing tumour/normal expression and using clustering analysis, the number of groups found to have the lowest Davis-Bouldin index (5 clusters, 1.17) were used to set a threshold for K-means clustering (Figure 5). Hierarchical clustering of 5 separate groups’ revealed separation between the five groups and KEGG pathway analysis of each subgroup was performed (Supplementary table 6). In three of the clusters there were either only one or two samples found. There was no distinction between these clusters in terms of anatomical location, stage or tumour mutational burden.

**Figure 5:**
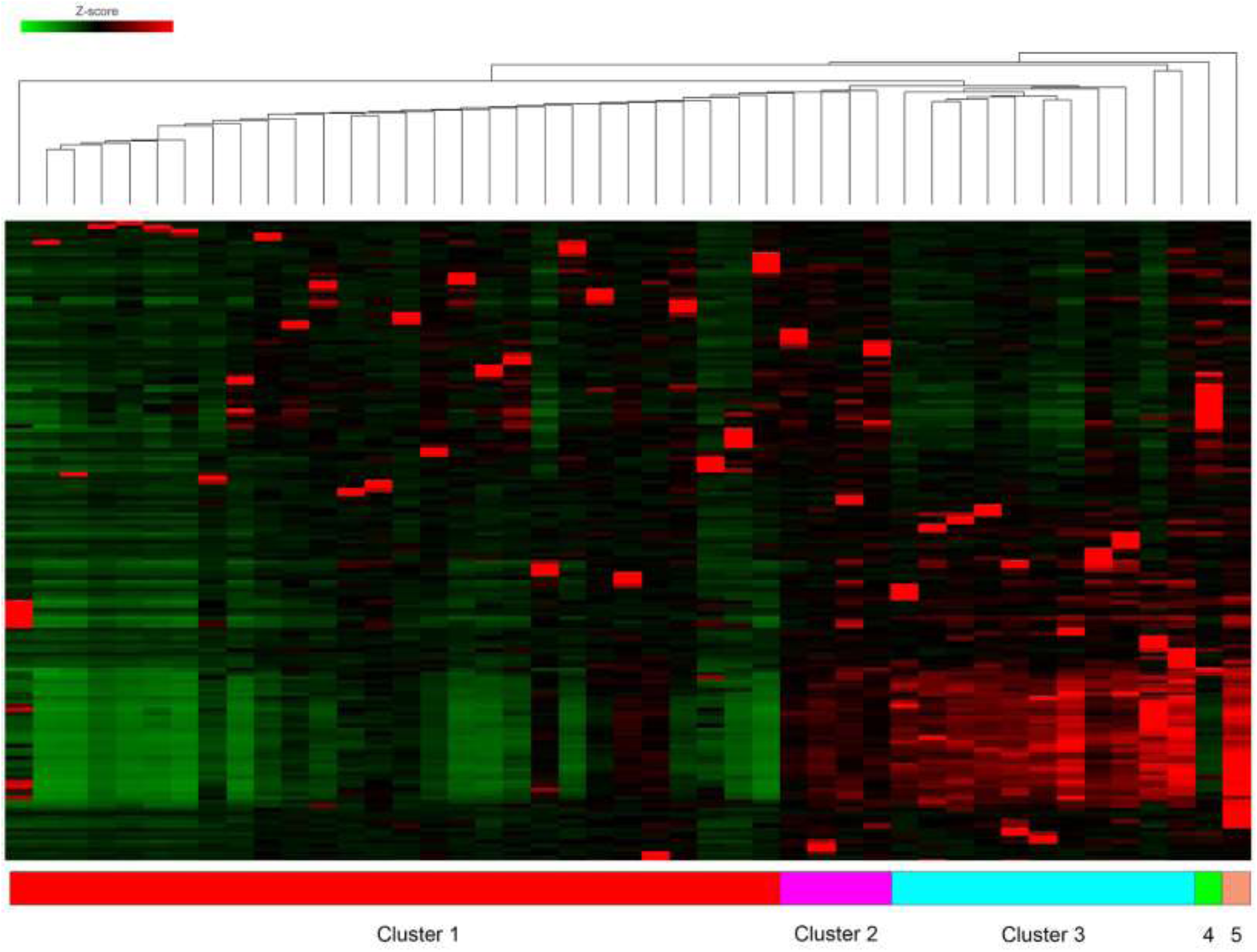
Hierarchical clustering plot of 100 most variably expressed genes in RNAseq data, demonstrating five separate clusters.

For subgroup one, an over-representation of pathways concerning inflammation and DNA repair was seen. For subgroups two and three no significant pathway over-representation was seen, possibly because these groups only had one sample within them. For subgroup four, multiple separate inflammatory pathways (mostly IL-17, Th1 and Th2 centric) were over-represented. Subgroup five had a number of interesting over-represented pathways, including reduced MHC presentation, Wnt/BMP signalling, TGFbeta signalling (via upregulated SMAD) and upregulated Hedgehog signalling.

### Pathway analysis

Single sample gene expression differences do not explain much of the context of disease processes, so we carried out a pathway gene expression analysis using the KEGG pathways of over-expressed genes to normal counts across the whole dataset. From this, we found a number of pathways of interest that were differentially expressed in colorectal cancer: the p53 signalling pathway (hsa41105, p=2.24×10–53, FDR_p_=1.06×10–51), NF-kappa-B signalling pathway (hsa040605, p=1.75×10–47, FDR_p_=4.95×10–46), and the ‘colorectal cancer’ pathway (hsa03030, p=2.06×10–41, FDR_p_=5.41×10–41) were all over-expressed in this cohort of patients.

A number of other pathways of interest (but not of direct relevance to colorectal cancer) were over-expressed, including platinum drug resistance (hsa01524), the Cytosolic DNA-sensing pathway (a.k.a. cGAS-STING, hsa04623) and several involved with DNA repair (FA pathway hsa03460, DNA replication hsa03030, NER, hsa03420).

## CMS/CRIS

Two classifiers for transcriptional subtypes in colorectal cancer have been identified (the Consensus Molecular Subtype (CMS) and the CRC Intrinsic Subtypes (CRIS) subtype (44, 45)), which reflect the disease biology of the tumour and have been linked with prognosis. These subtypes are derived from pre-existing molecular data by various computational methods to discover transcriptionally distinct groups within colorectal cancer. CMS and CRIS classifiers were generated for all tumours (Figure 6). Of the 54 sequenced tumours, the CMS classifier grouped the samples as follows: CMS 1 = 13/54, CMS2=9/54, CMS3=8/54, CMS4=12/54 and NA=3/54. For the CRIS classifier there were CRIS-A=9/54, CRIS-B=7/54, CRIS-C=12/54, CRIS-D=8/54, CRIS-E=5/54 and NA=4/54.

**Figure 6:**
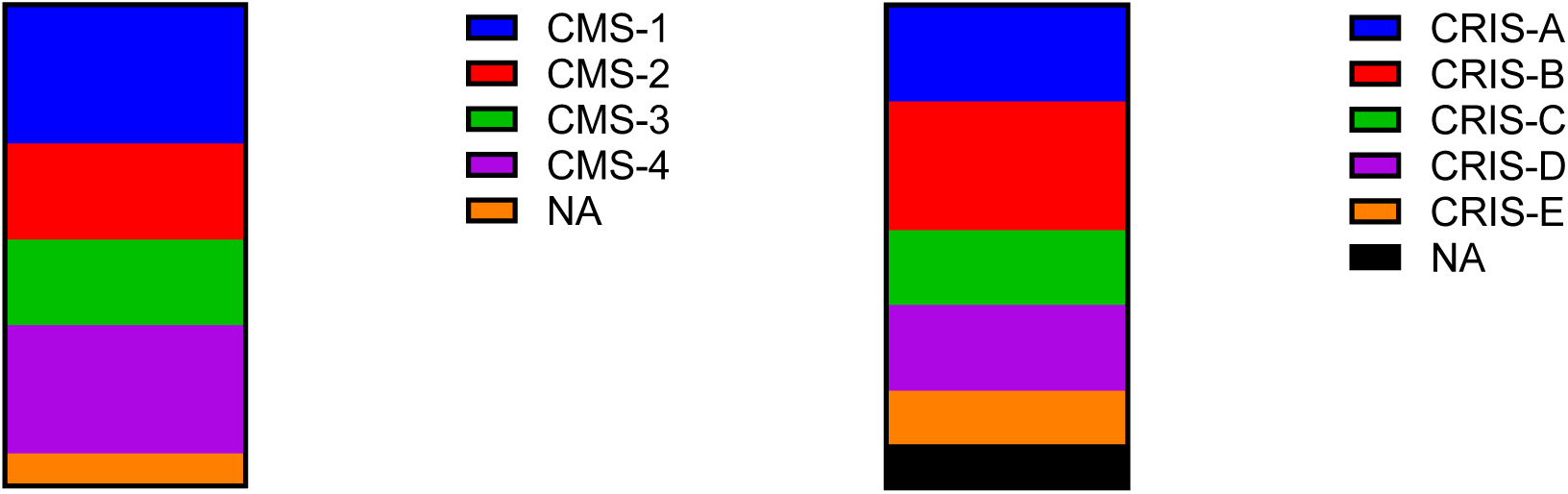
Graph of CMS calls (left) and CRIS calls (right) for dataset.

### CIRC

We have previously demonstrated the utility of the Coordinate Immune Response Cluster (CIRC) (30) as a Th1-centric RNA based signature in predicting Class I & II MHC immunovisibility (beyond TMB) in order to target with immunomodulatory drugs. Average of expression Z-score for the 28 genes in the CIRC was calculated for each tumour sample, with the lowest CIRC score being −0.56 and the maximum 3.17. In total, 12/54 samples had CIRC > 0 suggesting immunovisibility.

### Cell deconvolution using RNAseq

Immune infiltration estimation using cell type deconvolution by CIBERSORT (31)(Table 3) was performed on 3’ RNAseq data. This demonstrated a rich and varied immune infiltration within the colorectal cancers studied. The predominant cell type was CD4+ memory (resting) T-cell, followed by M2 macrophages, CD8+ T-cells, M0 macrophages then activated mast cells. There did not seem to be any correlation with purity estimates of the samples as determined by WGS.

**Table 3:** CIBERSORT classification of immune cells.

| Cell type | Score |
| --- | --- |
| T cells CD4 memory resting | 24.2 |
| Macrophages M2 | 11.9 |
| T cells CD8 | 11.3 |
| Macrophages M0 | 10.6 |
| Mast cells activated | 8.3 |
| B cells memory | 7.6 |
| NK cells activated | 7.5 |
| B cells naive | 5.5 |
| Dendritic cells activated | 4.7 |
| Plasma cells | 4.2 |
| T cells follicular helper | 4.1 |
| Neutrophils | 2.9 |
| Macrophages M1 | 2.7 |
| NK cells resting | 1.2 |
| T cells CD4 naive | 1.2 |
| T cells regulatory (Tregs) | 0.8 |
| Monocytes | 0.8 |
| Dendritic cells resting | 0.8 |
| T cells CD4 memory activated | 0.4 |
| Mast cells resting | 0.3 |
| Eosinophils | 0.09 |
| T cells gamma delta | 0 |

### RNA signature for hypermutation

In order to see whether a RNA based signature for hypermutation could be developed from RNA-seq data, gene-centric gene expression was processed using BioSigner (Bioconductor) using a threshold of >20 mutations/Mb in the WGS data (in order to develop a clear signature as >50% of hypermutant samples were near to the classical 10 mutations/Mb cut-off). Using 250 iterations of the algorithm, we attempted to generate Random Forest (RF), Partial Least Squares Discriminant Analysis (PLSDA) and Support Vector Machines (SVM) models of gene expression for hypermutant samples. We found that no stable model could be generated, however this could be a consequence of the relatively few numbers of hypermutant samples.

### Correlation between drug mutations database and druggable mutations

In order to ascertain the possibility of actionable targets from the mutations observed in the dataset, we entered a list of protein coding mutations found in at least one sample to the Drug Interaction Database (http://www.dgidb.org). Potential drug targets were observed for the genes – *APC, TP53, KRAS, FBXW7, ATM, PIK3CA, ARID1A, KMT2A, PTEN, SMARCA4, IDH1* and *RRM2B* (supplementary table 7). Also, 17/54 (32%) of patients exceeded the 10 mutations/Mb threshold for potential benefit for treatment with PD-1/PD-L1 therapy.

Utilising the OpenTarget platform (http://www.opentargets.org), which takes lists of mutations and functionally characterises them into drug targets, the 50 top genes from each tool for driver ranking (MutSigCV2, Intogen, dNdScv, Funseq2) were aggregated and input into the system (due to a limit of 200); after duplicate filtering this left 123 genes of interest. OpenTargets demonstrated significant enrichment for GI and epithelial tract cancers of all subtypes (Supplementary table 7). Also, significantly enriched pathways were seen in classical cancer pathways but also Interferon signalling, phagocytosis and Class I MHC signalling. Of the identified druggable genes, for small molecule agents, 8/123 had clinical precedence, 50/123 discovery precedence, and 49/123 were predicted to be tractable. Among antibody-based agents, 3/123 had clinical precedence, 68/123 had high tractable confidence and 83/123 had mid-low tractable confidence.

## CONCLUSIONS

The use of clinical grade whole genome sequencing in this study has allowed us to identify known and novel driver mutations that are potentially druggable based on the current state of knowledge. Our study demonstrated the known driver mutations seen in colorectal cancer such as *APC, KRAS, BRAF* and *PIK3CA* (5), but also more novel mutations that would potentially be targetable by molecular agents. For instance, we detected *KIT* mutations that would potentially be targeted by the tyrosine kinase inhibitor imatinib (46), offering a therapeutic option not available to these patients.

We also identified and validated several interesting potential driver mutations by frequency within our cohort. Recurrent mutations were seen in *KMT2C*, which codes for lysine methyltransferase-2C. These mutations have typically been seen in leukaemia and other blood malignancies but other more recent studies have demonstrated that these mutations occur amongst a wide variety of other cancers (47) and are targetable by inhibitors of KMT2C function. Mutations were also seen in *ATM* (targetable with ATM kinase inhibitors (48)), *IDH1* (targetable with the small molecular inhibitor of *IDH1*, Ivosidenib (49)) and *SMARCA4* (targetable with CDK4/6 inhibitors) (50). We attempted to identify new driver mutations as well as validate existing drivers using validated calling algorithms, however only *APC* was consistently enriched across all four callers in our study, once again emphasising the predominant Wnt signalling driven nature of colorectal cancer. The recurrent nature of *HLA-A* mutations (which were not validated by Sanger sequencing) in our cohort is interesting, as it is seen infrequently across all cancers (51), and could potentially represent a mechanism of immune invasion in a subset of cancers.

Recurrent alterations in genome structure, in the form of structural variants, copy number aberrations or gene fusions have also been highlighted as a potential target for therapy. For instance the *FGFR2/3* fusion seen in approximately 40% of cholangiocarcinoma is a target for the drug pemagatinib (52). Our study has shown several recurrent copy number variations or structural variations but also a number of unique “private” variations that may be targetable. For instance, we observed potential fusions between *BRAF* and *DLG1* (which may be targetable by BRAF kinase inhibition (53)) and between *ERBB2* and *HAP1* (which may be targetable by lapatinib (54)).

Tumour immunotherapy, using a combination of anti-PD1 and/or anti-CTLA4 therapy has been shown to have a survival benefit across multiple tumour types (55), especially when stratified to patients with high tumour mutational burden (TMB).

TMB correlates directly with neoepitope production and thus immunovisibility of the tumour. A threshold of 10 mutations per megabase of sequence has been suggested as a cut-off threshold sufficient for benefit for immunotherapy (56). Our study has shown that up to 20% of patients with colorectal cancer reach this threshold, which is higher (16%) than previously reported (5). This may be because whole genome sequencing provides a more comprehensive detection of mutations compared to other strategies, and also because of variations in how TMB is calculated.

We have carried out a variety of analyses of the RNA data derived from our samples. Surprisingly, the pathway analysis demonstrated findings of potential clinical utility, for instance, the presence of Kegg pathway hsa01524 (Platinum resistance). Oxaliplatin is commonly given in adjuvant chemotherapeutic treatment in colorectal cancer and resistance remains a problem (57), especially on the background of toxicity that leads to peripheral neuropathy. Interestingly, we have shown that the most frequent transcriptomic subtype within our dataset is CMS4, which is associated (44) with a worse prognosis (also seen in our dataset) and a more aggressive phenotype mainly due to the presence of fibroblasts which act as “malignant stroma”. The low numbers of accurate classification of our samples may represent a weakness of 3’ RNAseq (although we have previously used this technique without issue) or inherent weaknesses in the CMS classifier when a low tumour content heterogenous tumour sample undergoes sequencing (58). We have also demonstrated by cell deconvolution a rich and varied immune infiltration with the predominant cell types being CD4+ memory and CD8+ cells, however M2 macrophages are seen in most tumours. M2 macrophages are known as “repair” macrophages that decrease inflammation and promote tissue repair (59). If this is indeed the case it highlights an intriguing future path of research in colorectal cancer.

The CIRC classifier, which we have previously used to highlight immunovisibility (30) in cancer demonstrates that a proportion of samples have immunovisibility beyond those expected by high TMB.

In an era of personalised medicine, we have attempted to utilise current drug databases (DGIDb (60) and OpenTarget (61)) in order to identify targets for personalised medicine therapy. All patients had mutations within their tumour that were potentially “druggable” allowing their recruitment into a current or planned clinical trial. This is an exciting finding, as it gives a potential route of treatment for patients with metastatic disease, however the majority of these trials are phase one in nature and thus are not conclusively demonstrated to be active in colorectal cancer, or indeed in the targeted genomic alteration outside of pre-clinical models.

In conclusion, we have demonstrated the utility of standardised clinical grade WGS at detecting both new biological insights into colorectal cancer as well as targets for therapy. WGS has the advantage of breadth and depth of coverage but comes at the cost of expense; this is likely to drop significantly as technologies improve. A particular disadvantage in the clinical setting is the need for access to fresh-frozen tumour material in order to perform whole genome sequencing to the highest quality. The use of 3’ RNA seq allows a cost-effective way to further enrich the data returned by these assays and may be useful for future studies. The UK government has recently recommissioned Genomics England to sequence five million genomes over the next decade and we suggest based on our results that whole genome sequencing should be considered standard of care for colorectal cancer. We additionally suggest that RNA sequencing should be utilised as standard of care due to the additional insights it gives into tumour biology.

## Data Availability

Data will be made available on acceptance of the manuscript on EGA

---

**Supplementary Figure 1.**
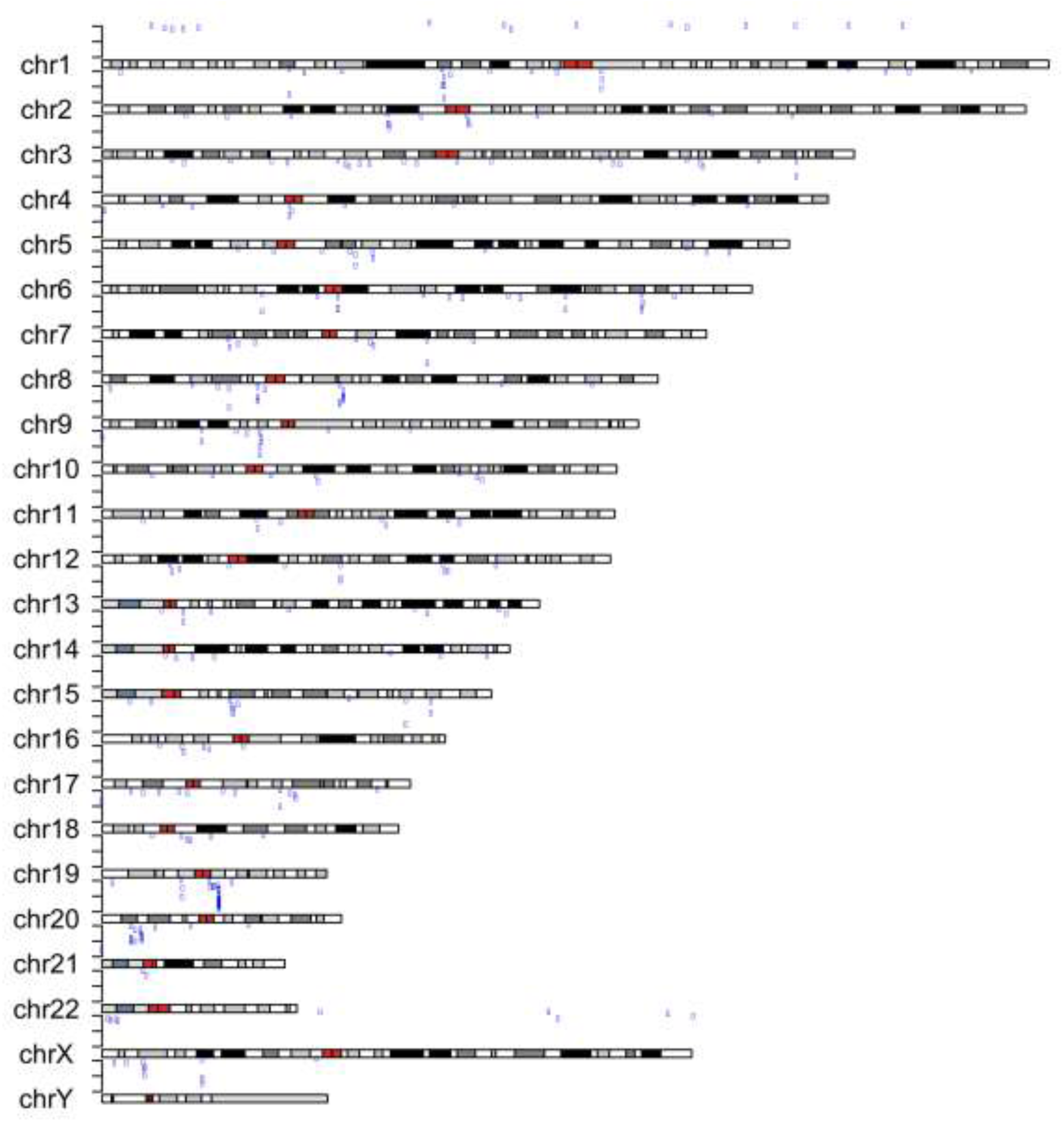
– Kataegis plot across whole genome for all samples. Regions of kataegis shown by blue lines above chromosomal plots.

## Notes

Funding: This study was supported by grants from the Academy of Medical Sciences (ref 102732/Z/13/Z) and the Wellcome Trust (ref 102732/Z/13/Z). ADB is currently supported by a Cancer Research UK Advanced Clinician Scientist award (ref C31641/A23923)

Conflicts of interest: AB has received travel costs and honoraria from Illumina Inc., Oxford Nanopore, Ono Pharm and Bristol Myers Squibb. MH, MV, ZK, JB, MR are employees of Illumina, a public company that develops and markets systems for genetic analysis.

### Competing Interest Statement

AB has received travel costs and honoraria from Illumina Inc., Oxford Nanopore, Ono Pharm and Bristol Myers Squibb. MH, MV, ZK, JB, MR are employees of Illumina, a public company that develops and markets systems for genetic analysis.

### Funding Statement

This study was supported by grants from the Academy of Medical Sciences (ref 102732/Z/13/Z) and the Wellcome Trust (ref 102732/Z/13/Z). ADB is currently supported by a Cancer Research UK Advanced Clinician Scientist award (ref C31641/A23923)

